# Modulating the Human Gut Microbiome and Health Markers through Kombucha Consumption: A Controlled Clinical Study

**DOI:** 10.1101/2024.07.01.24309793

**Authors:** Gertrude Ecklu-Mensah, Rachel Miller, Maria Gjerstad Maseng, Vienna Hawes, Denise Hinz, Cheryl Kim, Jack Gilbert

## Abstract

Fermented foods are surging in popularity globally due to their links to metabolic health and the gut microbiome. However, direct clinical evidence for the health claims is lacking. Here, we describe an eight-week clinical trial that explored the effects of a kombucha supplement in healthy individuals consuming a Western diet, randomized into the kombucha ( *n* = 16) or control (*n* = 8) group. We collected longitudinal stool and blood samples to profile the human microbiome and inflammation markers. Paired analysis between baseline and end of intervention time points within kombucha or control groups revealed increases in fasting insulin and in HOMA-IR in the kombucha group whereas reductions in HDL cholesterol were associated with the control group. Shotgun metagenomic analysis revealed the relative abundance of *Weizmannia*, a kombucha-associated probiotic to be overrepresented in consumers at the end of the intervention. Short-term kombucha intervention induced modest impacts on human gut microbiome composition and biochemical parameters.

**Highlights:**

- There is a global rise in popularity of fermented foods due to their links to metabolic health. However, empirical evidence supporting the health conferring benefits is lacking.
- This eight-week randomized controlled clinical trial tested the effect of kombucha on gut microbiome and inflammation in healthy participants consuming a Western diet.
- Biochemical indicators and inflammation markers were not different between the control and intervention groups at the end of the intervention.
- Kombucha-associated probiotic, *Weizmannia* and short chain fatty acids synthesizing microbes were enriched in consumers after 4 weeks of kombucha intervention.

**Graphical abstract:** 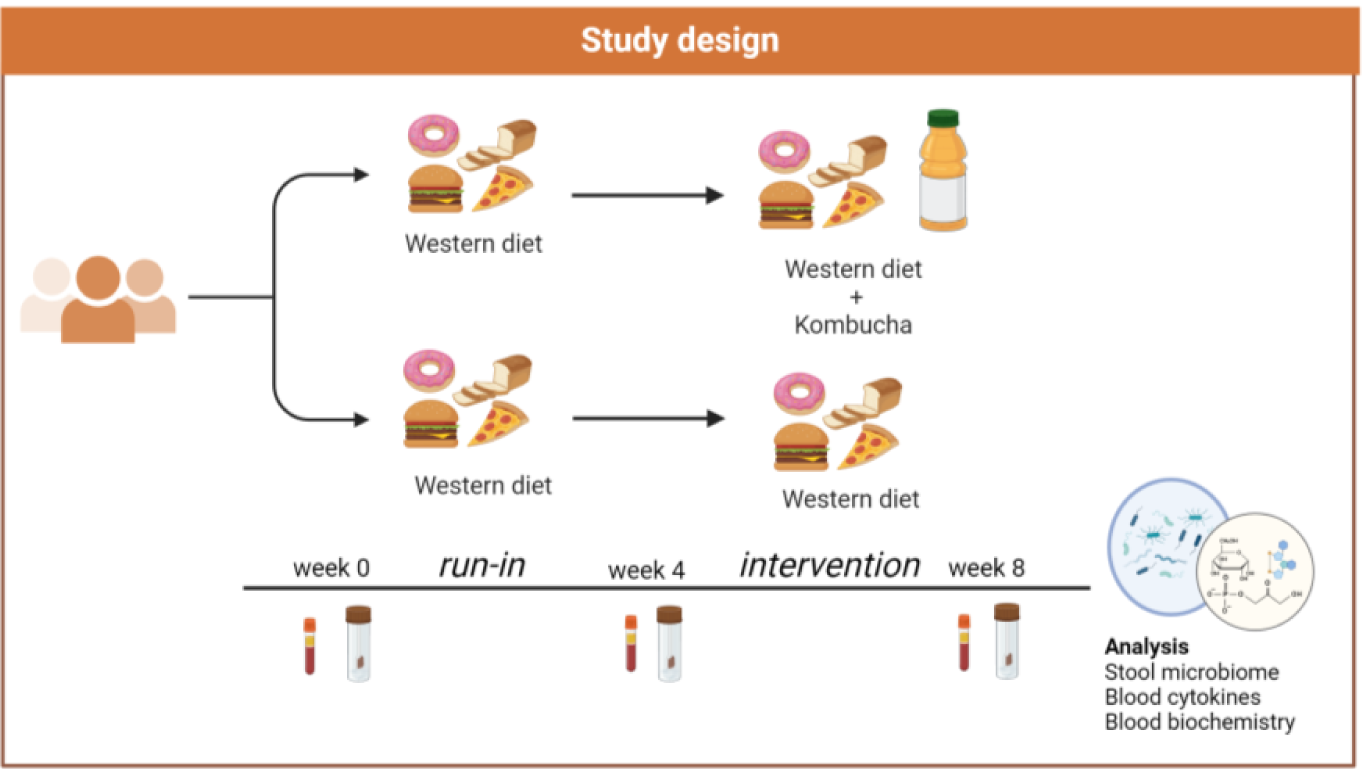

## 1. Introduction

Dietary strategies are recognized as significant modulators of the composition and function of the gut microbiome, which, in turn, can influence various aspects of human physiology, including metabolism. This interplay offers therapeutic promise for addressing diet-driven metabolic disorders (Asnicar et al., 2021; Mensah et al., 2024). Fermented foods containing bioactive compounds have garnered widespread interest and increasing consumer demand, particularly among modernized, industrialized populations, primarily driven by their perceived health-promoting benefits to minimize the impact of western diets (Dimidi et al., 2019; Hill et al., 2023; Marco et al., 2021; Mukherjee et al., 2024; Staudacher & Nevin, 2019). Observational and limited human dietary intervention studies have indicated associations between the consumption of diets rich in fermented foods and reduced disease risk, enhanced longevity, and improved quality of life (Bellikci-Koyu et al., 2019; González et al., 2019; Hill et al., 2023; Marco et al., 2017, 2021). For example, a systematic review and meta-analysis on the effect of fermented dairy products on cardiometabolic health revealed an association between fermented milk consumption and a reduced risk of type 2 diabetes (Gille et al., 2018). Also, a 2021 human dietary intervention study demonstrated that high consumption of fermented food increased microbiome diversity and correlated with improvements in several serum markers of inflammation, indicating that potential probiotic-like microorganisms in fermented foods may confer health benefits to humans (Wastyk et al., 2021).

While dairy-based fermented foods have been more extensively studied for their health benefits (Şanlier et al., 2019; van de Wouw et al., 2020), there is a relative lack of intervention studies on the specific health benefits of other fermented foods such as fermented tea beverages.

Kombucha is a non-alcoholic or low-alcohol tea-based beverage enriched with prebiotic compounds, acetic- and lactic-acid bacteria and yeast (Dimidi et al., 2019; Villarreal-Soto et al., 2020). Interactions between bacteria and yeast species can lead to the generation of a wide variety of metabolites with functional bioactivities, such as organic acids, vitamins, and phenolic compounds, which could potentially influence the gut microbiome and overall health (Cardoso et al., 2020; Mensah et al., 2024; Mukherjee et al., 2024; Villarreal-Soto et al., 2020). Several studies in animal models have suggested that intake of microbially-rich kombucha may offer potential therapeutic benefits, such as anti-hyperglycemic, anti-oxidant and anti-inflammatory effects, and changes in the gut microbiota that are associated with improved metabolic health (Cardoso et al., 2020; Jung et al., 2019; Lee et al., 2019; Morales, 2020; Moreira et al., 2022; Wang et al., 2021; Xu et al., 2022). For instance, Jung et al demonstrated that increased relative abundance of fecal *Lactobacillus* and a concurrent reduction in *Allobacullum, Turibacter and Clostridium* relative abundances correlated with suppression of liver fat accumulation and the mitigation of nonalcoholic fatty liver disease following kombucha administration in experimental mice (Jung et al., 2019). In another study, a four-week kombucha intervention in type 2 diabetes-induced mice reduced hyperglycemia and improved type 2 diabetes outcome through gut microbiota changes resulting in elevated proportional abundance of short chain fatty acids producing bacteria and decreased abundance of pathogenic bacteria (Xu et al., 2022).

Although, the mechanisms underpinning the observed effects have not been rigorously investigated, they are likely to occur through multiple inter-connected processes that include microbial food metabolites, the gut microbial ecosystem, and the host.

While promising results have been observed in animal models, direct evidence supporting the health conferring immune and metabolic health benefits of kombucha in human population remain sparse. Only two recently published controlled clinical trials have described the health benefits of kombucha consumption in human participants (Atkinson et al., 2023; Mendelson et al., 2023). For instance, in the Mendelson and colleagues study, consumption of kombucha for four weeks in adults with type 2 diabetes mellitus led to a significant fasting blood glucose reduction in comparison with baseline, a finding not observed in the placebo treated group.

However, none of the studies have directly evaluated the effects of kombucha intake on shifts in the gut microbiome. To drive the development of microbiota-directed interventions, human studies are necessary to understand the translational significance of pre-clinical findings and the potential benefit of kombucha on gut microbiome composition and overall human health.

Herein, we investigated the impact of regular consumption of kombucha for four weeks on markers of inflammation and composition of the gut microbiome of human adult participants consuming Western diets using metagenomic sequencing, biochemical and immune profiling. We showed that the kombucha supplement did not alter biochemical and inflammation profiles as a whole. Subtle changes in microbiota composition, driven by the enrichment of SCFA producing microbes including the kombucha-associated probiotic, *Weizmannia coagulans* were observed post-kombucha intervention. Our results reflect the relatively small number of participants, short study duration and the extensive inter-individual heterogeneity.

## 2. Materials and methods

### 2.1. Ethics statement

The research and related activities involving human subjects were approved by the Institutional Review Board at University of California, San Diego (IRB# 210823). All research, experiments and data analysis were performed in accordance with the University of California guidelines and regulations. All participants provided informed written consent prior to enrollment and all research was performed in accordance with the federal guidelines and regulations and the Declaration of Helsinki.

### 2.2. Study design

This was an eight-week, randomized clinical controlled trial where individuals were assigned 2:1 to either a non-kombucha control group or a kombucha intervention group. There were 3 time points in the study, namely, time points 1 (baseline), 2 (four weeks from baseline) and 3 (eight weeks from baseline). Participants were randomized using a random number generator in Excel by a biostatistician who was not involved in the intervention or data collection. The first four weeks after time point 1 data completion, participants consumed a beige/Western diet (i.e. low-fiber, low polyphenol diet), thereafter, participants randomized into the intervention group consumed one bottle (two servings) of a 16 oz commercial kombucha daily for four weeks in addition to their beige diet, whereas control-randomized participants maintained their beige diet intake. During each study visit at time points 1, 2 and 3, fasting blood was drawn, while anthropometrics data, stool samples, questionnaires and three-day food records were collected. The study was designed as exploratory with the primary endpoint as changes in microbiota diversity after kombucha intervention, while secondary endpoints included changes in biochemical and immune markers (IL6, IL10, CRP). The trial has been registered at ClinicalTrials.gov, identifier: NCT06484504.

### 2.3. Eligibility and recruitment

Free-living participants were recruited from the San Diego County through online and paper advertisements as well as social media platforms (Facebook, LinkedIn, Twitter) and emails sent to the University of California (UC) San Diego community. We assessed 60 participants for eligibility where they answered questions on medical history, lifestyle habits, age, sex, antibiotic use, bowel movement, and diet, followed by a clinic visit for eligible participants. The primary inclusion criteria included the following: healthy adults aged 21-55 years, females must be non-menopausal, BMI between 18-29.9. Participants were excluded if they had a history of gastrointestinal surgery, diabetes mellitus on medications, or other serious medical condition, such as chronic hepatic or renal disease, bleeding disorder, congestive heart disease, chronic diarrhea disorders, myocardial infarction, coronary artery bypass graft, angioplasty within 6 months prior to screening, current diagnosis of uncontrolled hypertension (defined as systolic BP ≥160mmHg, diastolic BP ≥ 95mmHg), Other exclusion criteria included use of antibiotics or laxatives within the last three months, using prebiotics, probiotics, and/or any fiber supplements regularly and allergy or sensitivity to kombucha. Thirty participants passed the screening and were enrolled in the study.

### 2.4. Product description

The investigated product used was a 16 oz of a commercial kombucha beverage, which according to the manufacturer contained a propriety blend of three immune boosting probiotic strains. The concentrations of the probiotics *Bacillus coagulans* GBI-306086, *S*. *boulardii* and *Lactobacillus* bacterium were one billion, four billion and four billion organisms respectively. Other naturally occurring components in the product were lactic acid (100mg), acetic acid (75mg), glucuronic acid (1400mg) and gluconic acid (650mg). Participants were instructed to take half portion of one bottle of the 16 oz kombucha twice a day, every day for the four weeks of intervention along with their Western diet.

### 2.5. Sample collection and processing

Participants were requested to fast for at least 10 hours prior to each clinic visit at the Altman Clinical Translational Research Institute, UC San Diego. Blood samples were collected from the antecubital vein into SST (serum separator tube) and EDTA vacutainers (Becton Dickinson, Franklin Lakes, NJ, USA), span at 1,200xg for 10 minutes for serum and plasma respectively, aliquoted, and stored at −80°C for downstream analysis. Blood collected for metabolic parameters were analyzed within two hours of collection at the CLIA-certified Center for Advanced Laboratory Medicine at UC San Diego. A wall-mounted stadiometer and an electronic scale were used to measure height and weight, respectively. Body mass index was calculated by dividing the weight (kg) by the height squared (m^2^). Waist circumference was measured midway between the lowest rib and the iliac crest using an anthropometric nonelastic tape, blood pressure, pulse and respiration were measured using the Mindray Accutorr 7 equipment. Participants were provided sterile stool swab sampling kits with instructions on sample collection. Stool swab samples were collected at three different time points (1, 2, 3). Specifically, within three days prior to each study visit at time points 1 (baseline), 2 (four weeks) and 3 (eight weeks), participants collected three sets of stool swab samples which were preserved in ethanol and brought to each clinic visit. Stools samples were submitted at each clinic visit and then transferred to −80°C storage in the laboratory. At the end of the study, each participant was expected to submit nine stool samples in total.

### 2.6. Laboratory analysis

#### 2.6.1. Blood biochemistry

Standard metabolic indicators were analyzed using Roche cobas® 8000 modular analyzer following standard procedures within two hours of blood collection. Homeostatic Model Assessment for Insulin Resistance (HOMA) index was calculated as insulin level (µU/ml) × glucose level (mmol/l)/22.5. Additional biochemical parameters evaluated included lipid levels (total cholesterol, HDL cholesterol, LDL cholesterol, triglyceride), creatinine level, and estimated glomerular filtration rate (eGFR).

#### 2.6.2. Inflammation markers

Data on inflammation markers were generated at the La Jolla Institute for Immunology. Briefly, the quantifications of IL-6, IL-10, and C- reactive protein (CRP) from serum were determined using bead-based LEGENDplex immunoassays as described by the manufacturer’s protocols (BioLegend, San Diego, CA). LEGENDplex assays were measured using a BD FACSCanto^TM^ II flow cytometer (BD Biosciences), and data analyzed using LEGENDplex software (BioLegend) and R version 4.1.1.

#### 2.6.3. Metagenomic analysis

Nucleic acid extractions were performed at the UC San Diego Microbiome Core using previously published protocols (Marotz et al., 2021). Briefly fecal samples were randomly sorted, transferred to 96-well extraction plates and DNA was extracted using the MagMAX Microbiome Ultra Nucleic Acid Isolation Kit (Thermo Fisher Scientific, USA) and automated on KingFisher Flex robots (Thermo Fisher Scientific, USA). Blank controls and mock controls (ZymoBiomics) were included per extraction plate, which were carried through all downstream processing steps. Purified DNA was quantified using a PicoGreen fluorescence assay (Thermo Fisher Scientific, USA) and metagenomic libraries were prepared with KAPA HyperPlus kits (Roche Diagnostics, USA) according to the manufacturer’s instructions. Sequencing was performed on the Illumina NovaSeq 6000 sequencing platform with paired-end 150 bp cycles at the Institute for Genomic Medicine (IGM), UC San Diego. Raw metagenomic sequences were quality filtered and trimmed using fastp v 0.22 (S. Chen et al., 2018) and sequence reads mapping to the human genome and PhiX reads were filtered using minimap2 version 2.22 (Li, 2018, 2021) and converted to FASTQ format using SAMtools version 1.15.1. (Danecek et al., 2021). Reads were aligned against the Web of Life reference database (Zhu et al., 2019) using Bowtie2 (Langmead & Salzberg, 2012) and classified with the Woltka pipeline (Zhu Qiyun et al., 2022) to generate a table of Operational Genomic Units (OGUs). Sequences matching the human reference genome and genomes with less than 0.5% coverage per sample and features with less than ten reads in the entire dataset were removed.

### 2.7. Statistical analyses

The biochemical and anthropometric data of the study participants were summarized using descriptive statistics, with continuous variables presented as median and interquartile range, while categorical variables were presented as number and percentages. Significant changes were evaluated from time point 1 to end of intervention (time point 3) by a paired Wilcoxon test (within groups) and unpaired Wilcoxon test (between control vs. kombucha groups). All microbiome samples were rarified to 2,959,909 reads, retaining 210/241 samples and alpha diversity measures quantified as Observed OGUs and Shannon Index were calculated from the OGU table using phyloseq (McMurdie & Holmes, 2013). Beta diversity was determined using Bray Curtis dissimilarity, weighted and unweighted UniFrac distances (Lozupone & Knight, 2005) generated in phyloseq and statistical differences were assessed with permutational multivariate analysis of variance (PERMANOVA) using the adonis2 function in the Vegan package. To assess longitudinal distance from baseline, sample dissimilarity between baseline and other time points for kombucha and control samples were calculated using the miaTime R package v0.1.15. Relative abundances at the phylum and species level were calculated using the unrarefied OGU table and presented as percentages. To determine differentially abundant features between control and intervention groups over time, the unrarefied OGU table was binned at the species level and used in the analysis of compositions of microbiomes with bias correction (ANCOMBC) (Lin & Peddada, 2020). Sex, BMI, and age were added as covariates in the ANCOMBC formula. All statistical tests were two-sided and *p* < 0.05 was deemed statistically significant. Adjustments for multiple comparisons (*q* values) were applied as indicated. Data analysis and figures were generated in R version 4.1.1.

## 3. Results

### 3.1. Free living participants successfully completed kombucha intervention

To determine the effect of kombucha consumption on free-living adults on Western diets, we recruited participants for an eight-week randomized, controlled study. Of over 100 individuals initially expressing interest, 30 eligible individuals maintained interest after learning about all the study requirements and agreed to be enrolled in the trial. Participants were randomized (2:1) into the kombucha group (n =20) or control group (n = 10). The primary outcome was change in microbiome composition at the end of the kombucha intervention period, while the secondary outcome was change in metabolic and immune markers after kombucha intervention.

At the end of the trial, we had two participants, one in each group, taking antibiotics along the study time, one individual with higher HbA1c (> 5.7%), two participants lost to follow up without specific reasons and one individual who experienced bloating during the intervention and had to withdraw from the study (Figure 1). These participants were excluded from the analysis, and hence the number of participants was reduced to 24 (16 intervention, 8 controls), obtained from 11 male and 13 female participants. All participants provided stool and blood samples at baseline (timepoint 1), four weeks prior to the intervention (timepoint 2) and at the end of the four-week intervention period (timepoint 3) in which participants in the kombucha group took two servings of kombucha (16 oz in total per day).

**Figure 1.**
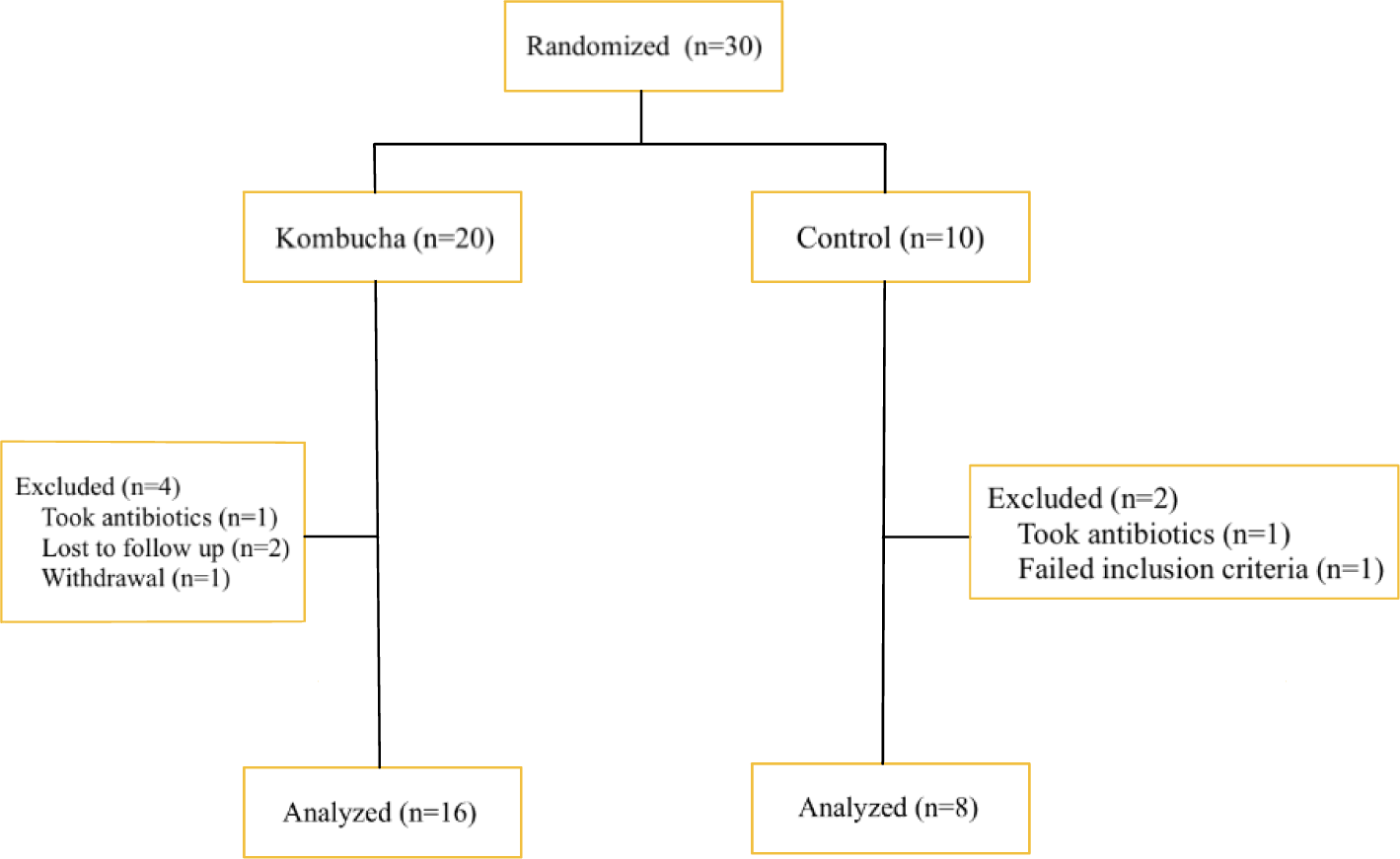
Flow diagram of study

### 3.2. Kombucha intervention did not influence on anthropometric, metabolic or immune parameters

We analyzed the anthropometric measurements and as shown in Table 1, we did not find any statistically significant difference between the control and the kombucha groups at any time point of the study. The secondary outcome of the study was change in metabolic health parameters in participants consuming kombucha from time point 1(baseline) to end (time point 3) of intervention. There were no significant changes in diastolic blood pressure, systolic blood pressure, fasting glucose levels, triglycerides, HDL cholesterol, or waist circumference over time between groups (unpaired Wilcoxon test) except for baseline cholesterol levels where control participants had significantly higher levels (unpaired Wilcoxon test; p=0.035; Table 1). Fasting insulin and LDL cholesterol, elevated levels of which are associated with deteriorating health also did not differ between time points 1 and 3 between groups (unpaired Wilcoxon test; Table 1). Within the kombucha group (*n* = 16), significant increases were observed for fasting insulin levels (paired Wilcoxon test; p=0.021) and HOMA-IR (paired Wilcoxon test; p=0.021) over time. A significant decrease in HDL cholesterol levels (paired Wilcoxon test; p=0.042) and marginal increase in creatinine levels (paired Wilcoxon test; p=0.050) were observed for the control group (n=8) over time.

**Table 1.** Biochemical and anthropometric characteristics of participants.

| Variable | Baseline |  | Final |  | P-1 | P-2 | P-3 |
| --- | --- | --- | --- | --- | --- | --- | --- |
|  | Kombucha (n=16) | Control (n=8) | Kombucha (n=16) | Control (n=8) |  |  |  |
| BMI (kg/m<sup>2</sup>) | 23.7(4.28) | 23.63(4.61) | 23.59(3.93) | 23.63(4.98) | 0.78 | 0.84 | 0.29 |
| Age(years) | 25.5(6.5) | 26 (9.5) | 25.5(6.5) | 26(9.5) | 1 | na | na |
| Anion gap (mmol/L) | 9(2) | 8.5(1.5) | 10(1.25) | 10(1.5) | 0.34 | 0.4 | 0.43 |
| Bicarbonate (mmol/L) | 27(3.25) | 26.5(2.5) | 26(3) | 26(1.5) | 0.78 | 0.35 | 0.07 |
| BUN (mg/dL) | 10.5(4.75) | 10.5(7) | 13.5(6) | 9(1.75) | 0.98 | 0.11 | 0.57 |
| Calcium (mg/dL) | 9.7(0.83) | 9.65(0.23) | 9.6(0.33) | 9.6(0.3) | 1 | 0.5 | 0.18 |
| Chloride (mmol/L) | 103(1.25) | 103.5(1.25) | 103(2) | 103(1.5) | 0.85 | 0.34 | 0.58 |
| Cholesterol (mg/dL) | 155(25.75) | 186.5(34.75) | 163(25.75) | 195(64.75) | **0.035** | 0.83 | 0.33 |
| Creatinine (mg/dL) | 0.755(0.17) | 0.82(0.11) | 0.76(0.16) | 0.85(0.19) | 0.81 | **0.05** | 0.88 |
| Diastolic BP (mmHg) | 70(14.38) | 66.75(15.5) | 72.25(16) | 71(8.38) | 1 | 0.57 | 0.92 |
| Systolic BP (mmHg) | 116.25(12.13) | 111(17.5) | 118(12.13) | 114.5(11.88) | 0.43 | 0.94 | 0.81 |
| EGFR (mL/min) | 61(0) | 61(0) | 61(0) | 61(0) | 0.96 | 1 | 0.2 |
| Fasting insulin (uu/mL) | 6.15(2.98) | 7.4(5.38) | 9.3(4.83) | 8.6(2.2) | 0.65 | 0.95 | **0.021** |
| Glucose (mg/dL) | 90.5(12.25) | 92(3.5) | 91.5(7.25) | 91.5(11.5) | 0.67 | 0.67 | 0.47 |
| HbA1c (%) | 5.05(0.3) | 5.1(0.3) | 5.3(0.33) | 5.2(0.15) | 0.95 | 0.92 | 0.09 |
| HDL-C (mg/dL) | 56.5(13.75) | 63(20.5) | 58(14.25) | 54(21.25) | 0.48 | **0.042** | 0.78 |
| Height (cm) | 172.2(13.59) | 178.73(25.65) | 172.03(13.94) | 179.18(24.79) | 0.44 | 0.93 | 0.16 |
| Hip (cm) | 96.45(10.1) | 103.5(12.31) | 96.36(8.51) | 101.2(7.03) | 0.3 | 0.84 | 0.33 |
| HOMA-IR | 1.392(0.89) | 1.69(1.27) | 2.01(1.29) | 1.82(0.53) | 0.6 | 0.84 | **0.021** |
| LDL cal (mg/dL) | 91.5(15.5) | 103(24.5) | 96.5(25.25) | 111(47.5) | 0.19 | 0.58 | 0.31 |
| Non HDL-C (mg/dL) | 100.5(14.5) | 121.5(38.25) | 109.5(31.75) | 131.5(53.75) | 0.12 | 0.4 | 0.38 |
| Potassium (mmol/L) | 4.35(0.475) | 4.3(0.4) | 4.35(0.33) | 4.25(0.23) | 1 | 0.32 | 0.38 |
| Pulse | 63.5(10.38) | 69.25(12.13) | 63.5(16) | 65.5(14.63) | 0.52 | 0.4 | 0.86 |
| Respiration | 16(0.5) | 16(1) | 15.75(3) | 15.75(2) | 0.77 | 0.39 | 0.6 |
| Sex, female (%) | 10(62.5) | 3(37.5) | 10(62.5) | 3(37.5) | na | na | na |
| Sodium (mmol/L) | 116.25(12.13) | 140.5(1.75) | 139(2) | 138.5(2.25) | 0.46 | 0.32 | 0.14 |
| Temperature (oC) | 36.65(0.23) | 36.65(0.15) | 36.675(0.23) | 36.6(0.11) | 0.9 | 0.29 | 0.88 |
| Triglyceride (mg/dL) | 59(25.25) | 69.5(20) | 57(38) | 72(29.25) | 0.52 | 0.67 | 0.66 |
| Waist (cm) | 84.675(7.36) | 83.75(14.65) | 81.55(8.33) | 81.875(13.09) | 0.58 | 0.64 | 0.27 |
| Weight (kg) | 71.125(13.29) | 78.35(18.58) | 71.225(12.14) | 77.8(18.69) | 0.41 | 0.55 | 0.92 |

Data are shown as median with interquartile range. Sex is shown as number of participants (with the percentage in brackets). BP, blood pressure; DBP, BUN, Blood urea nitrogen; HDL-C, high-density lipoprotein cholesterol; LDL-C, low-density lipoprotein cholesterol; BMI, body mass index. *p*-values, as determined by 2-sided Wilcoxon test, between or within groups are shown. P-1, *p* value for control baseline versus kombucha baseline; P-2, *p* value for control baseline vs control final; P-3, *p* value for kombucha baseline vs kombucha final; na, not analyzed.

Despite the lack of generalized changes in several metabolic health parameters in the kombucha group relative to the control group, we wondered whether there were detectable changes in aspects of the immune system of participants in response to kombucha consumption. Studies, notably in animal models suggest that kombucha has immunomodulatory properties, positively influencing inflammation cytokines expression (Costa et al., 2023; Haghmorad et al., 2021; Wang et al., 2021). Therefore, we reasoned that changes in markers of inflammation could be an indication that kombucha supplementation could beneficially impact the immune system of participants. The sera of participants were analyzed for three different circulating cytokines (IL10, IL6 and CRP). No significant changes in immune features from baseline to end of intervention within participants were observed in either the kombucha or control groups (Figure 2). Direct comparison of control and kombucha arms at the end of the intervention also did not reveal any differences indicating that the kombucha intervention did not influence inflammation in our cohort.

**Figure 2.**
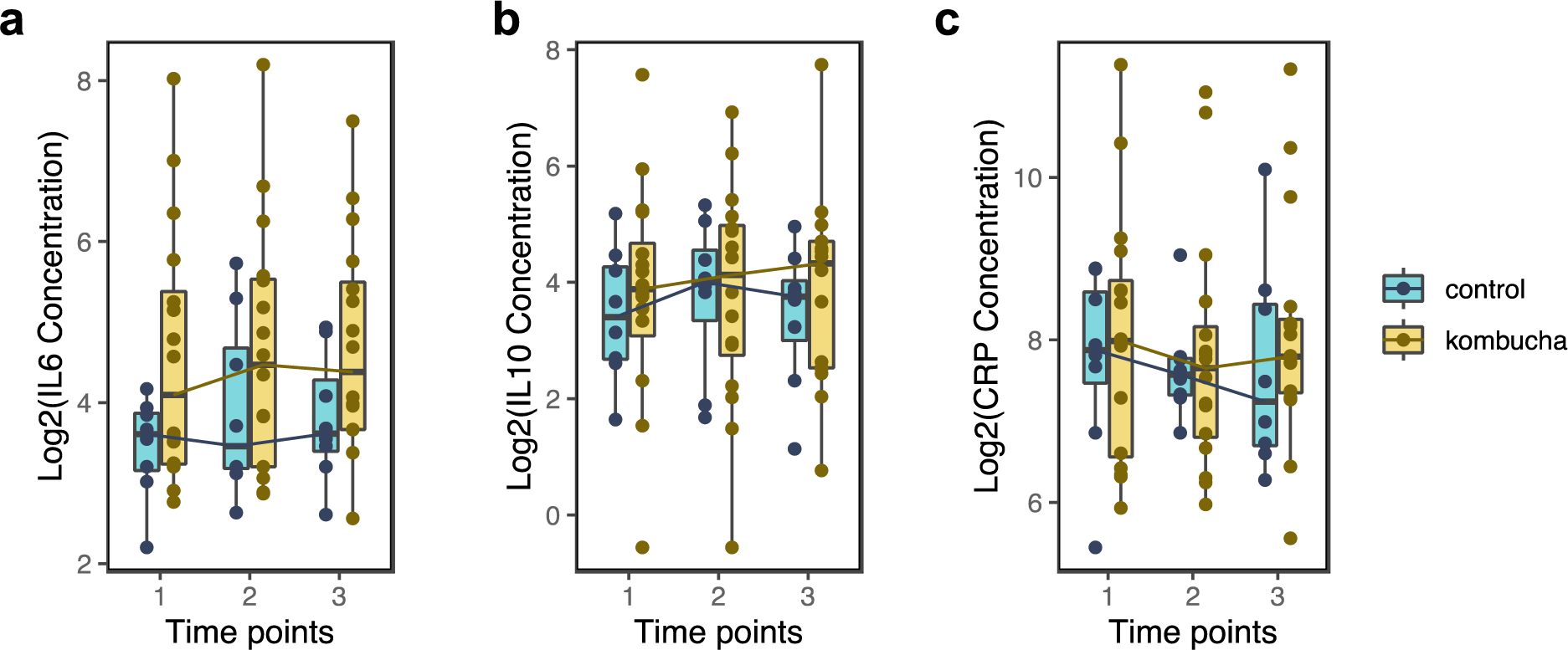
Differences in serum inflammation marker concentrations between the kombucha and control groups over time. IL6, interleukin 6; IL10, interleukin10; CRP, c-reactive protein. There were no significant differences within or between groups at each time point as assessed by Wilcoxon test (p>0.05).

### 3.3. Kombucha consumption marginally shifts gut microbiota profiles

We hypothesized that any effect of kombucha consumption would be captured in a microbiome-induced shift and demonstrable in stool samples. To characterize the effect of kombucha consumption on the microbiota of adults, fecal samples were subjected to shotgun sequencing and Operational Genomic Units (OGUs) were assigned by the Web of Life reference database (Zhu et al., 2019). After quality filtering, a total of 2,369,069,908 reads with a median of 9,647,881 reads and 2,128 taxa were generated from 200 samples (2-3 samples per 3 time points) from 24 unique participants. Since each participant collected 2-3 fecal samples from consecutive days at each time points, these replicates were merged for analyses. Merged replicates generated a median of 28,353,277 sequences from 72 samples, which was rarefied to equal depth of 2,959,909 reads per sample for diversity measures.

Compositional analysis showed that the gut microbiota of participants in the kombucha and control groups across all timepoints was dominated by 5 major phyla, namely Bacteroidota (47%), Firmicutes_A (30.9 %), Actinobacteriota (5.18 %), Proteobacteria (7.53%) and Verrucomicrobiota (6.00%), contributing over 96% of the total composition (Supplementary figure 1a). The top 10 prevalent genera were *Bacteroides, Phocaeicola, Akkermansia, Escherichia, Alistipes, Prevotella, Bifidobacterium, Gemmiger, Agathobacter* and *Faecalibacterium* (Supplementary figure 1b). In figure 3a which captures the relative abundance of the top 20 species in each group, *Prevotella* was associated with the kombucha group, whereas *Escherichia* was observed in the control group.

**Figure 3.**
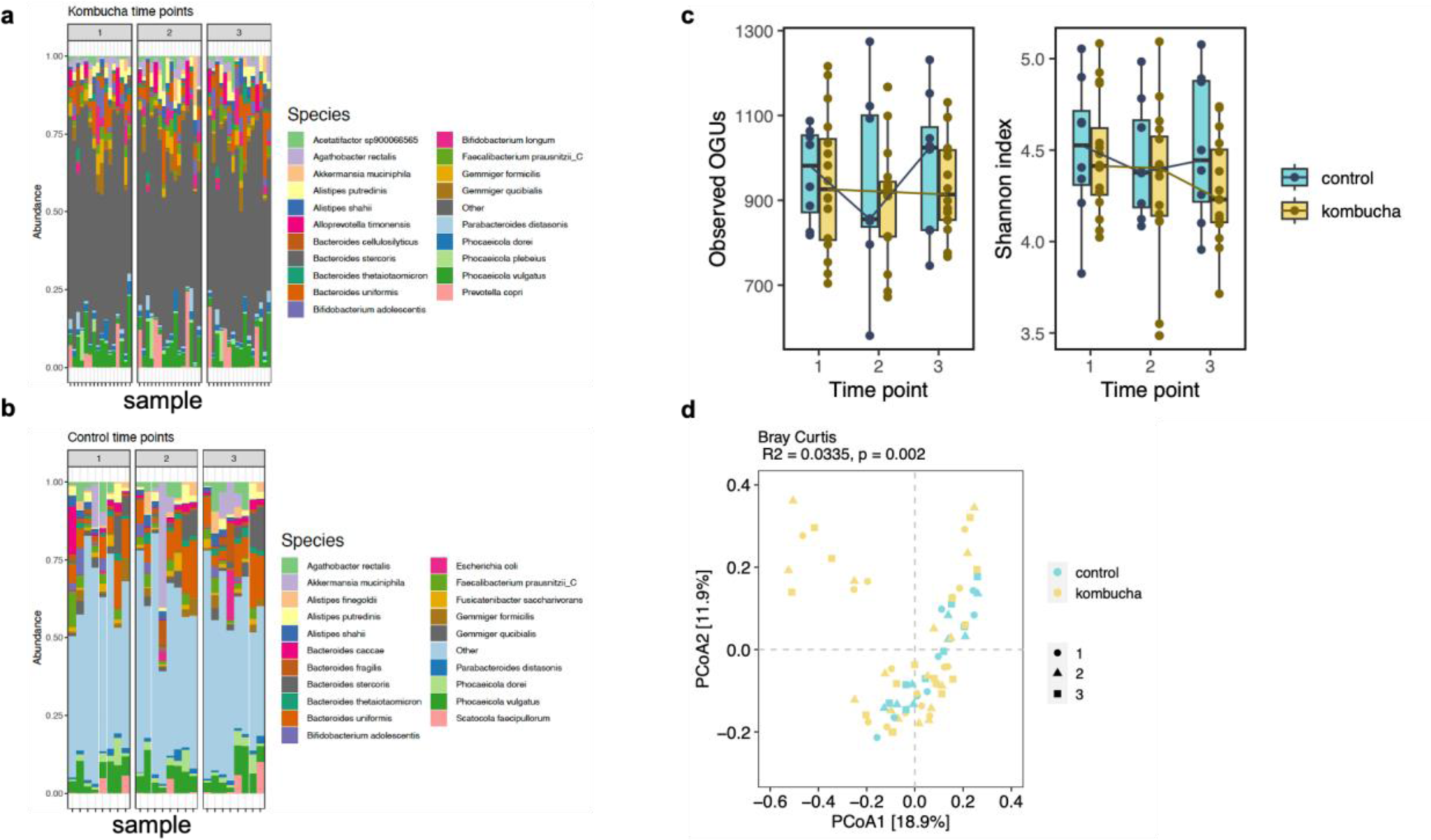
Changes in gut microbiome between kombucha and control groups over time. a. The relative abundance of top 20 bacterial species in gut microbiome of control samples. b. The relative abundance of top 20 bacterial species in gut microbiome of intervention samples. Each vertical bar represents an individual sample. c. Observed OGUs and Shannon diversity for control and intervention groups at each time point. Shannon diversity significantly decreased between time point 1 and time point 3 for the kombucha group (Wilcoxon test). Boxplots denote the interquartile range (IQR) between the first and third quartiles, and the horizontal line defines the median. d. Principal coordinate analysis (PCoA) of gut microbiota community structure measured by Bray–Curtis dissimilarity and calculated for OGU level composition. Composition of gut microbiota differed significantly between control and kombucha group with all time points included. Significance testing was performed with permutational analysis of variance (PERMANOVA; permutation *n* = 999). OGUs, operational genomic units.

Alpha (α) diversity was measured using the Shannon Index and Observed OGUs with host id as a random effect. There was no significant difference in Observed OGUs between the intervention and control groups over time (Fig. 3b; LME; p>0.05). However, we noted a significant reduction in the Shannon Index from time point 1 to the end of study (LME; p=0.029) and a trend towards significance from timepoint 1 to time point 2 (LME; p=0.05). At the end of intervention, kombucha and control groups did not differ significantly in any of the measured α- diversity indices (time point 3; p>0.05; unpaired Wilcoxon test).

Within groups, Shannon Index decreased from timepoint 1 to time point 2 (LME; p=0.043) and also from time point 1 to timepoint 3 (LME; p=0.0058) with host id as random effect in the kombucha group, however significance remained only for time point 1 vs time point 3 (q= 0.016) after multiple testing correction. The observed decline in the Shannon Index suggests a potential marked shift among certain microbial species at the end of intervention, resulting in altered evenness in species distribution and decreased overall gut microbiota diversity. By contrast, there was no differences in microbiota α-diversity (Shannon and Observed) over time within the control group.

Next, we calculated 3 different beta diversity measures (Fig. 3c; Supplementary figure 1c&d) and found significant differences in overall composition between kombucha and control groups with all time points included (unweighted uniFrac, PERMANOVA, p=0.007; weighted uniFrac, PERMANOVA, p=0.006, Bray Curtis, PERMANOVA, p=0.002). To determine whether the microbiota composition of participants changed over the course of the study, we calculated the Bray Curtis dissimilarity from each participant’s microbiota at time point 1 to all other time points and found no significant changes within or between groups. At the end of the intervention, the microbiota composition of the kombucha group did not differ from the control group (PERMANOVA, *p* > 0.05). Together, our results suggest that while study participant accounted for most of the variation, time and kombucha intervention had little effect on total microbiota community structure dissimilarity.

To examine whether specific microbial species were significantly different between the kombucha and control groups at the end of intervention, we ran the ANCOMBC algorithm for compositional differential abundance testing, which identifies genomic features characterizing the differences between two or more biological conditions (Lin & Peddada, 2020). Results from this analysis shown in figure 4, revealed that the microbiota of participants in the kombucha group were enriched in the relative proportions of several species including a kombucha-associated bacteria, *Weizmannia coagulans* compared with those of the control group at the end of intervention (Fig. 4a). Similarly, *Weizmannia coagulans* was differentially abundant among other species in the kombucha group at the end of intervention compared with baseline or time point 2 (Fig. 4b-d). Our observed results indicated that kombucha intervention resulted in detectable changes in the gut microbiota of consumers. Finally, no statistically significant evidence was found for correlations between OGUs that differed following 4 weeks of kombucha consumption and the significant biochemical parameters (fasting insulin, creatinine, HDL cholesterol, HOMA-IR) using Spearman’s correlation coefficient.

**Figure 4.**
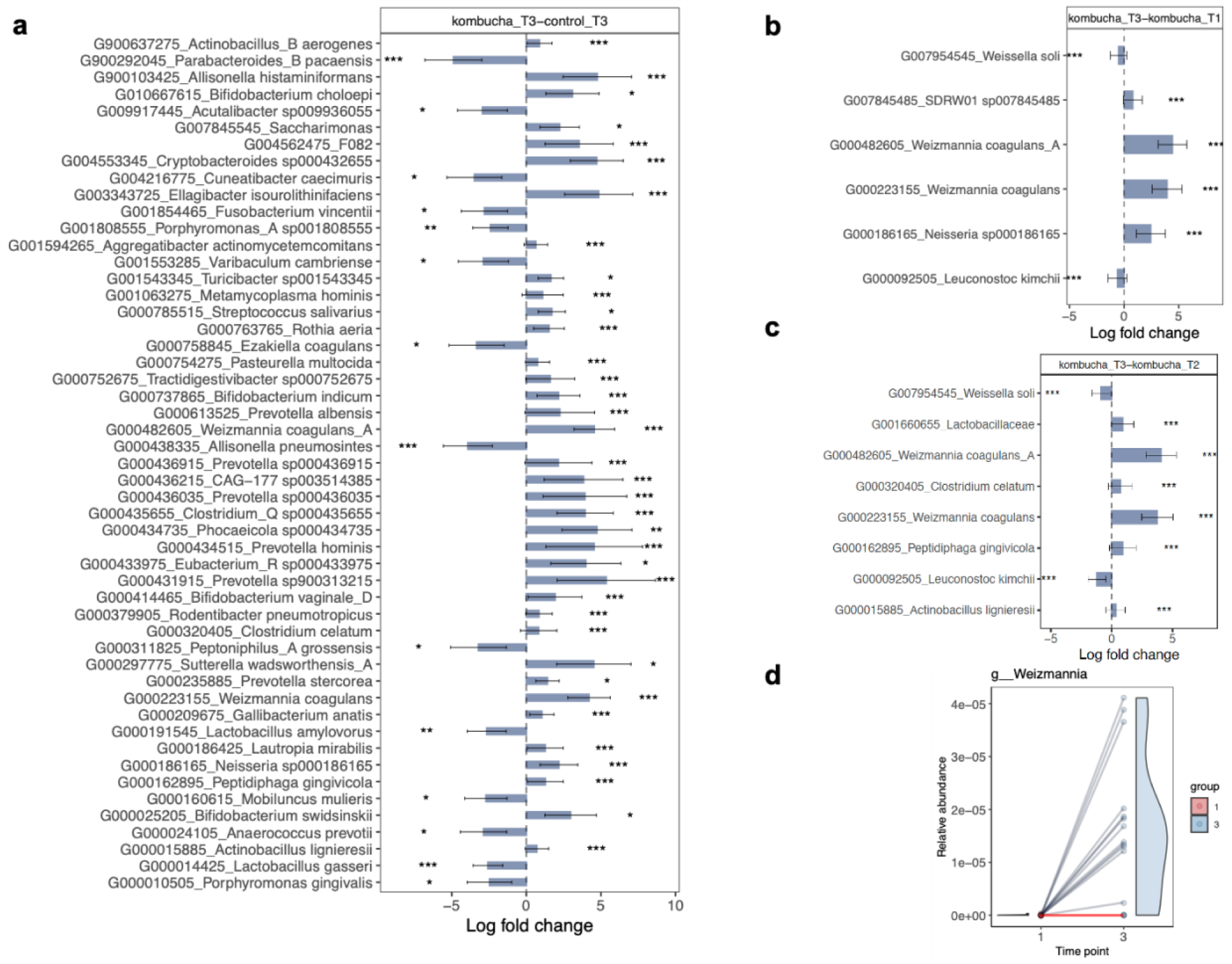
Microbial features associated with kombucha intervention. Differentially abundant features at the species level in gut microbiome samples in a. control (*n* = 8) vs kombucha (*n* = 16) groups at the end of intervention (time point 3). b. Kombucha microbiome samples before (time point 1) and after intervention. c. Kombucha microbiome samples at time point 2 pre-intervention and after intervention. Bars represent the log-fold-change (effect size) of individual bacterial features showing significant effect sizes (*q* < 0.05). A positive log-fold-change indicates that a feature is more abundant in participants in the post-treatment kombucha group (Kombucha_T3), and a negative log-fold-change indicates a higher abundance in participants in the comparison group (control_T3, Kombucha_T1, Kombucha_T2). Lines represent standard error derived from the ANCOMBC (Analysis of compositions of microbiomes with bias correction) model. Changes in the relative abundance of all members of the *Weizmannia* genus pre- (T1) and post- (T3) intervention in the kombucha group.

## 4. Discussion

Here, we describe a randomized controlled human study examining the effects of a daily portion of kombucha dietary supplementation in free-living healthy individuals consuming a Western diet, combining high dimensional, longitudinal microbiome and markers of systemic inflammation profiling. We did not observe changes in biochemical parameters between kombucha consumers and controls, a finding consistent with previous randomized clinical trials in healthy cohorts (Walsh et al., 2023; Wastyk et al., 2021). Paired analysis between baseline and end of intervention for kombucha or control groups however, revealed increases in fasting insulin and HOMA-IR in kombucha group whereas reductions in HDL cholesterol were associated with the control group. Levels of circulating markers of inflammation were not significantly different between kombucha consumers and controls or within groups over time, suggesting that kombucha supplementation may have little impact on inflammation in healthy participants. Analysis of shotgun metagenomic sequencing data revealed that overall microbial diversity did not differ between kombucha consumers and controls, though marked differences in microbiota community composition and individualized microbiomes were observed. Within the kombucha group, microbiota diversity decreased modestly between baseline and end of intervention. Further, we found kombucha-associated *Weizmannia coagulans* to be enriched in consumers, suggesting that consumption of kombucha may trigger shifts in the gut environment. Taken together, our findings suggest that in our healthy cohort, only minor effects of kombucha intervention on human gut microbiota structure and biochemical parameters are observed, which may be attributed to relatively small number of participants and the extensive inter-individual variability.

Several studies have indicated that dietary interventions may cause profound changes in microbiota structure (David et al., 2014; Oliver et al., 2021; von Schwartzenberg et al., 2021; Wastyk et al., 2021). While in this study, a four week-daily kombucha consumption resulted in a minimal decrease in microbiota diversity, a finding which contrasts with the increased microbial diversity frequently associated with individuals consuming fermented foods (Galena et al., 2022; Wastyk et al., 2021). The lack of large-scale microbial changes could partly be attributed to study factors, such as sample size or short duration of the study which might not have provided sufficient time for microbiota reshaping. Our study participants maintained a low fiber Western diet and it is possible that kombucha intervention on a background of Western diet has limited impact on microbial diversity in humans. Most studies have utilized a diverse mix of fermented foods (Berding et al., 2022; Guse et al., 2023; Taylor et al., 2020; Wastyk et al., 2021) in contrast to our study that relied on a single fermented food supplement approach. Furthermore, existing studies vary in the quantity as well as the type of fermented food and duration of consumption, fermented food matrices, study designs, participant characteristics and health outcomes targeted. For example, increasing consumption of different fermented foods to six servings per day over 10 weeks in an intervention study elicited changes in gut microbial diversity (Wastyk et al., 2021). By contrast, participants consumed 2 servings of kombucha for four weeks, suggesting that in our short intervention study, this fermented food threshold or magnitude may have been insufficient to induce broad microbial remodeling as observed previously (Wastyk et al., 2021). Nevertheless, our findings are in line with other studies that have reported no changes to microbiota diversity as a result of fermented food intake in comparison with controls or pre-intervention state (Alvarez et al., 2020; Berding et al., 2022; Guse et al., 2023; Taylor et al., 2020; Walsh et al., 2023). For instance, a randomized, double-blind, controlled intervention study evaluated the effect of daily consumption of two doses of a multi-strain fermented milk product for 4 weeks on the gut microbiome. The authors reported no changes in microbial alpha diversity after the intervention with the fermented product (Alvarez et al., 2020). Similarly, habitual consumption of plant based fermented foods did not alter alpha diversity in relation to non-consumers (Taylor et al., 2020).

While overall shifts in microbial diversity were unaffected, our study observed that the composition of the gut microbiome was impacted by kombucha supplementation. We detected significantly altered differential abundance of selected taxa between consumers and controls at the end of intervention, a pattern consistent with earlier studies (Galena et al., 2022; Guse et al., 2023; Taylor et al., 2020; Walsh et al., 2023; Wastyk et al., 2021). Specifically, we observed the enrichment of 36 species and the concurrent depletion of 14 taxa associated with kombucha consumers. We found the enrichment of short-chain fatty acids producing taxa including several species within the genus *Prevotella* and *Bifidobacterium*, *Eubacterium*, *Clostridium*, and *Ellagibacter isourolithinifaciens* and bacteria with less-favorable effects such as *Neiserria*.

Importantly, of the microbial species present in the kombucha brand consumed, *Weizmannia coagulans* (*Bacillus coagulans*) was more frequently detected in the gut microbiome samples of consumers. Indeed, within the kombucha group, changes in community composition post- kombucha intervention were largely driven by shifts in *Weizmania coagulans*. Additionally, we detected significant depletions in the relative abundance of *Parabacteroides*, *Allisonella*, *Lactobacillus* and *Fusobacterium* among others in the kombucha cohort.

The dominance of *Weizmannia coagulans* in the kombucha consumers indicates that within four weeks of kombucha administration, the gut microbiome composition is characterized by an increase in bacteria ingested from the product. *Weizmannia coagulans* (*Bacillus coagulans*) is a Gram-positive, lactic acid-producer, non-pathogenic and spore-forming bacterium, widely used as a commercial probiotic in foods (Konuray & Erginkaya, 2018; Maresca et al., 2024).

*Weizmannia coagulans* is a preferred probiotic due to its resistance to high temperature, high survival rate under harsh low-oxygen environments of the gastrointestinal tract, stomach acids and bile salts as well as food processing (Maresca et al., 2024; Tripathi & Giri, 2014).

*Weizmannia coagulans* has many known effects on digestive health, nutrient absorption, and human health through multiple mechanisms, including enzyme production, metabolite generation and modulation of the gut environment (Cao et al., 2020). Its remarkable characteristics may explain its potential colonization in the gut following four-week kombucha consumption. Most claims about functional and health promoting properties related to kombucha are due to the high content of phenolic compounds present in the beverage (Bellassoued et al., 2015; Dutta & Paul, 2019; Villarreal-Soto et al., 2019; Vīna et al., 2014). Certain strains of *Weizmannia coagulans* can restructure the gut microbiota by increasing the abundance of intestinal transforming ellagic acid bacteria such as *Ellagibacter isourolithinifaciens* to facilitate the bioavailability of polyphenols (P. Chen et al., 2016; Jin et al., 2022). In our study, we detected proportional increases of *Ellagibacter isourolithinifaciens*, after four weeks of kombucha consumption, known to be involved in dietary polyphenol metabolism in healthy individuals (Beltrán et al., 2018; García-Villalba et al., 2020). As kombucha is rich in polyphenols, we speculate that kombucha consumption may have promoted the abundance of *Ellagibacter isourolithinifaciens* in the gut of consumers, although we did not measure levels of polyphenols in our study. Altogether, it is unclear if the observed changes are sufficient to elicit noticeable health benefits in our healthy participants, especially as we did not detect any profound differences in the biochemical parameters measured here.

While fermented foods have been associated with anti-inflammatory effects (Bourrie et al., 2023; Marco et al., 2017; van de Wouw et al., 2020; Wastyk et al., 2021; Zhang et al., 2023), the absence of significant differences in the inflammatory profile of our study participants may be attributed to our limited sample size, short duration of the intervention and the quantity of fermented food consumed by participants. It is possible that other parameters of the immune system that are more malleable to dietary modulation in healthy individuals including white blood cell profile, T cells or a more comprehensive immune profiling (Menni et al., 2021; Wastyk et al., 2021) could provide detailed insights into the immunomodulatory capabilities of kombucha.

Our exploratory investigation focused on a limited number of healthy participants over a short duration, potentially constraining our ability to discern significant impacts on microbial, biochemical and inflammation profiles. Moreover, the modest sample size contributed to intra-individual variations that surpassed the inter-individual heterogeneity in gut microbiota composition. Without microbial metabolites data, we were unable to explore possible mediators in response to kombucha intake in healthy participants. While our study suggests that kombucha supplementation may have influenced compositional shifts in the microbiome, we acknowledge the presence of other influencing factors, such as host genetics or nondietary behaviors. Moving forward, larger-scale randomized studies conducted over extended periods within clinical cohorts combining biomarkers of immune, metabolic, and endocrine function with microbiome data are essential to thoroughly explore the potential therapeutic utility of microbially-rich kombucha beverages in optimizing health outcomes.

## 5. Conclusions

A short term kombucha dietary intervention in healthy participants differentially influenced the composition of gut microbiota, enriching several SCFA producing taxa. However, these compositional changes did not correspond to broad shifts in biochemical or inflammation profiles, at least over this short-term intervention. Applying multi-omic approaches on larger sample size with longer study duration are needed to delineate the impact of kombucha consumption on gut microbiota modulation and its connection to human health and disease outcomes.

## CRediT authorship contribution statement

**Gertrude Ecklu-Mensah:** Conceptualization, Methodology, Formal analysis, Data curation, Visualization, Project administration, Writing - original draft, Writing - review & editing. **Rachel Miller:** Methodology, Data curation, Writing - review & editing. **Maria Gjerstad Maseng:** Methodology, Formal analysis, Writing - review & editing. **Vienna Hawes:** Methodology, Writing - review & editing. **Denise Hinz:** Methodology, Data curation, Writing - review & editing. **Cheryl Kim:** Methodology, Data curation, Writing - review & editing. **Jack Gilbert:** Conceptualization, Methodology, Project administration, Supervision, Funding acquisition, Writing - review & editing.

## Data Availability

All data produced in the present work are contained in the manuscript

## Acknowledgments

The authors thank the study participants for their engagement and essential contribution to the study. We appreciate Joshua Tran, Davis Bone and Juston Jaco for providing support during study clinic visits and data entry and Dr. Sarah Allard for proof reading and feedback on the manuscript. Funding was provided by GT’s living foods LLC.

## Appendix A. Supplementary material

