## Supplemental Information for "Modulating the Human Gut Microbiome and Health Markers through Kombucha Consumption: A Controlled Clinical Study"

Supplementary Table 1. Biochemical and anthropometric characteristics of participants at time point 3

| | Final | | |
|---|---|---|---|
| **Variable** | **Kombucha (n=16)** | **Control (n=8)** | **P value** |
| BMI (kg/m2) | 23.59(3.93) | 23.63(4.98) | 0.88 |
| Age(years) | 25.5(6.5) | 26(9.5) | 1 |
| Anion gap (mmol/L) | 10(1.25) | 10(1.5) | 0.45 |
| Bicarbonate (mmol/L) | 26(3) | 26(1.5) | 1 |
| BUN (mg/dL) | 13.5(6) | 9(1.75) | 0.08 |
| Calcium (mg/dL) | 9.6(0.33) | 9.6(0.3) | 0.42 |
| Chloride (mmol/L) | 103(2) | 103(1.5) | 0.83 |
| Cholesterol (mg/dL) | 163(25.75) | 195(64.75) | 0.28 |
| Creatinine (mg/dL) | 0.76(0.16) | 0.85(0.19) | 0.48 |
| Diastolic BP (mmHg) | 72.25(16) | 71(8.38) | 0.95 |
| Systolic BP (mmHg) | 118(12.13) | 114.5(11.88) | 0.34 |
| EGFR (mL/min) | 61(0) | 61(0) | 0.78 |
| Fasting insulin (uu/mL) | 9.3(4.83) | 8.6(2.2) | 0.76 |
| Glucose (mg/dL) | 91.5(7.25) | 91.5(11.5) | 0.83 |
| HbA1c (%) | 5.3(0.33) | 5.2(0.15) | 0.35 |
| HDL-C (mg/dL) | 58(14.25) | 54(21.25) | 0.48 |
| Height (cm) | 172.03(13.94) | 179.18(24.79) | 0.37 |
| Hip (cm) | 96.36(8.51) | 101.2(7.03) | 0.12 |
| HOMA-IR | 2.01(1.29) | 1.82(0.53) | 0.88 |
| LDL cal (mg/dL) | 96.5(25.25) | 111(47.5) | 0.48 |
| Non HDL-C (mg/dL) | 109.5(31.75) | 131.5(53.75) | 0.26 |
| Potassium (mmol/lL) | 4.35(0.33) | 4.25(0.23) | 0.56 |
| Pulse | 63.5(16) | 65.5(14.63) | 0.5 |
| Respiration | 15.75(3) | 15.75(2) | 0.7 |
| Sex, female (%) | 10(62.5) | 3(37.5) | na |
| Sodium (mmol/L) | 139(2) | 138.5(2.25) | 0.78 |
| Temperature (oC) | 36.675(0.23) | 36.6(0.11) | 0.14 |
| Triglyceride (mg/dL) | 57(38) | 72(29.25) | 0.36 |
| Waist (cm) | 81.55(8.33) | 81.875(13.09) | 0.69 |
| Weight (kg) | 71.225(12.14) | 77.8(18.69) | 0.43 |

Data are shown as median with interquartile range. Sex is shown as number of participants (with the percentage in brackets). BP, blood pressure; DBP, BUN, Blood urea nitrogen; HDL-C, high-density lipoprotein cholesterol; LDL-C, low-density lipoprotein cholesterol; BMI, body mass index. *p*-values, as determined by 2-sided Wilcoxon test, between groups are shown.

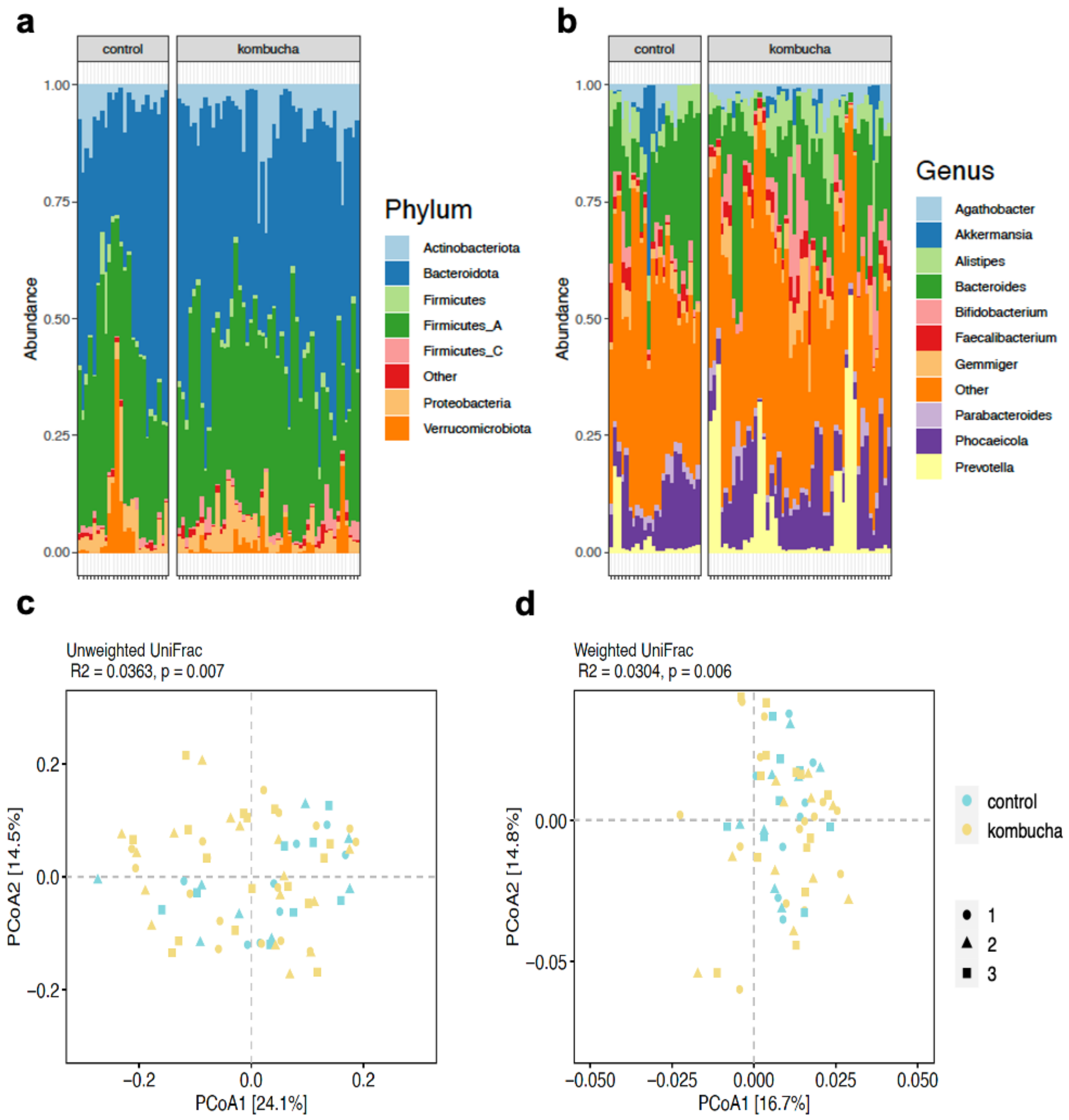


Supplementary Figure 1. Variation in gut microbiome between kombucha and control groups over time. a. The relative abundance of top seven bacterial phyla in gut microbiome of control and intervention samples. b. The relative abundance of top 10 bacterial genera in gut microbiome of control and intervention samples. Each vertical bar represents an individual sample. c. Principal coordinate analysis (PCoA) of gut microbiota community structure measured by Unweighted Unifrac distance and calculated for OGU level composition. d. Principal coordinate analysis (PCoA) of gut microbiota community structure measured by Weighted Unifrac distance and calculated for OGU level composition. Composition of gut microbiota differed significantly between control and kombucha group with all time points included. Significance testing was performed with permutational analysis of variance (PERMANOVA; permutation *n* = 999). OGUs, operational genomic units.
